# Comparison of Clinical Outcomes in Acute Coronary Syndrome Patients Who Undergo Percutaneous Coronary Intervention Using Different P2Y12 Inhibitors: A SMS Hospital Based Observational Study

**DOI:** 10.1101/2023.12.04.23299360

**Authors:** Raghuraj Swami, Sohan Kumar Sharma, Pradeep Meena

## Abstract

**Background:** To compare the safety, and efficacy of contemporary P2Y12 inhibitors, prescription rates, drug defaulter and switch over rates in acute coronary syndrome (ACS) patients following percutaneous coronary intervention (PCI).

**Methods:** in this prospective observational study we studied 195 ACS patients who underwent PCI in SMS Hospital, Jaipur. We compared prescription rates, bleeding, and major adverse cardiac events (MACEs: cardiac death, nonfatal myocardial infarction, or stroke) according to ticagrelor, prasugrel, or clopidogrel use.

**Results:** The prescription rates of ticagrelor, prasugrel, and clopidogrel were 29.5%, 4.3%, and 66.2% respectively. Bleeding occurred in total 6 patients (2.9%) out of which 3 patients (2.2%) in clopidogrel group, 1 patient (11.1%) in prasugrel group and 2 patients (3.2%) in ticagrelor group (p=0.29)), respectively, with higher incidence in ticagrelor and prasugrel users than in clopidogrel users but statistically not significant. After six month follow up, all-cause mortality was total 5 cases (2.4%) out of which 4 death (2.9%) occurred in clopidogrel group while 1 death (1.6%) occurred in ticagrelor group(p=0.76). no mortality was recorded in prasugrel group. Repeat myocardial infarction (Re MI) occurred in total 4 patients (1.9%) with 2 patients in clopidogrel and ticagrelor group each. A total 5 patients (2.4%) switch over to another p2y12 inhibitor, mostly from ticagrelor group(p=0.045). During this follow up period, a total of 4 patients (1.9%) were drug defaulter (0.622).

**Conclusions:** As clopidogrel based DAPT was prescribed more in our centre because of free institutional supply and found no significant difference in clinical outcomes, although slight increase in bleeding risk in ticagrelor or prasugrel based DAPT, demonstrate clopidogrel based DAPT regime can be preferably prescribed safely in our population. Drug defaulter rates in first six month following PCI are low in all groups but more switch over from ticagrelor group to other groups significantly shows importance of low cost and free institutional supply in drug adherence.

## INTRODUCTION

Dual antiplatelet therapy (DAPT), containing aspirin and a P2Y12 inhibitor, is recommended in acute coronary syndrome (ACS) patients undergoing percutaneous coronary intervention (PCI) to decrease atherothrombotic risk.^1-3^

But, clopidogrel has insufficient efficacy because its platelet inhibitory action is slow, modest, and very uneven.^2,3^

Because ticagrelor and prasugrel shows faster, stronger, and more consistent potency than does clopidogrel and have more favorable clinical benefits in randomized trials ^2-6^, they were recommended over clopidogrel in ACS patients undergoing PCI.^1^

The Prasugrel Versus Ticagrelor in Patients with Acute Myocardial Infarction Treated with Primary Percutaneous Coronary Intervention (PRAGUE-18) trial reported that in acute myocardial infarction (MI) patients who underwent PCI, both newer p2y12 inhibitors, ticagrelor and prasugrel had a comparable efficacy and safety.^7,8^

However, real-world data on contemporary P2Y12 inhibitors, especially which used in ACS patients undergoing PCI, are limited and inconsistent.^9-12^

Based on the reported association between contemporary P2Y12 inhibitors and outcomes, we will investigate the prescription rate, efficacy, and safety of contemporary P2Y12 inhibitors and also drug defaulter and switch over rates in ACS patients undergoing PCI in SMS hospital Jaipur.

## AIMS AND OBJECTIVES

Our primary objective was to compare the efficacy and safety of contemporary P2Y12 inhibitors in acute coronary syndrome (ACS) patients following percutaneous coronary intervention (PCI)

Our secondary objective were to compare prescription rates, adherence to the treatment and study various cause for drug defaulter or switch over to other P2Y12 inhibitor.

## MATERIALS AND METHODS

### Study Design

Hospital based observational, prospective study.

### Study Location

Department of cardiology, SMS Medical College and Hospital, Jaipur

### Study Duration

Until the sample size is reached with six month follow up.

### Sampling Size

Sample size of 210 patients of ACS who undergo PCI are required at 80% study power and alpha error of 0.05

### Sampling Procedure

Newly diagnosed 210 patients of ACS who underwent PCI and prescribed DAPT were included in the study.

### Inclusion Criteria

All patients diagnosed ACS undergoing PCI in SMS Hospital, Jaipur were included in the study after willful written and informed consent given by patient.

Patients aged ≥18 years, those with a confirmed final diagnosis of ACS, those who undergo PCI, and those prescribed aspirin and a P2Y12 inhibitor (ticagrelor, prasugrel, or clopidogrel) before PCI were included.

### Exclusion Criteria

Patients with aged <18 years, without follow-up, and who not given willful and written consent were excluded.

## METHODS

Patients with angina-like symptoms, presented to the emergency department/ cardiology opd of SMS hospital, Jaipur, diagnosed ACS were enrolled.

Before PCI, each patient was routinely receive antiplatelet agents, including aspirin 325 mg, and a P2Y12 inhibitor (ticagrelor 180 mg, prasugrel 60 mg, or clopidogrel 600 mg) followed by daily aspirin (75 mg) indefinitely and a P2Y12 inhibitor (ticagrelor 90 mg twice daily, prasugrel 10 mg once daily, or clopidogrel 75 mg once daily) for at least 1 year.

Prasugrel was not be prescribed in patients >75 years old, <60 kg in weight, or with prior stroke/transient ischemic attack.

The antiplatelet agents were chosen largely based on the discretion of individual cardiologists.

Unfractionated heparin (50-70 U/kg) was administered before or during PCI to maintain the activated clotting time at 250-300 seconds.

Angiotensin-converting enzyme inhibitors or angiotensin receptor blockers, platelet glycoprotein IIb/IIIa inhibitors, statins, and β-blockers were administered per the physician’s discretion.

All patients were undergo coronary angiography according to current standard procedural guidelines.

The interventional cardiologist determined the specific PCI technique and stent type for the coronary lesion. Standard care and medications for secondary prevention was provided to patients.

After discharge, all patients were prescribed medications and were followed up regularly (1- to 3-month intervals) by opd visits and telephonically. During follow-up, patients who developed angina-like symptoms undergo complete clinical evaluations.

If deemed necessary, patients received hospital care and revascularization. Information on clinical events was obtained from hospital records or via telephone contact with the patients’ relatives.

## STUDY END POINTS AND DEFINITIONS

The primary efficacy end point was incidence of major adverse cardiac events (MACEs; cardiac death, nonfatal MI, or stroke) during follow up.

Secondary end points were MACE components, all-cause death, non-cardiac death, any revascularization including repeat PCI (re-PCI), and CABG during follow-up.

All-cause death was defined as any case of death intra- or post-procedure; death was considered to be of a cardiac origin, unless a definite non-cardiac cause will be established.

Nonfatal MI was defined as recurrent symptoms with new electrocardiographic changes compatible with MI or cardiac marker levels at least twice the upper limit of normal.

Stroke was defined as a new, sudden, focal neurological deficit due to a presumed cerebrovascular cause that was not reversible within 24 hours and not due to a readily identifiable cause (e.g., tumors or seizures).

Any revascularization was defined as revascularization involving either the target or non-target vessels.

The primary safety end point were cumulative in-hospital bleeding events unrelated to coronary artery bypass grafting (CABG), as defined by the Thrombolysis in Myocardial Infarction bleeding criteria.^13^

Major bleeding included intracranial bleeding, clinically overt sign of hemorrhage, and a decline in hemoglobin level of ≥5 g/dL or in hematocrit levels of ≥15%.

Minor bleeding was defined as any bleeding requiring medical intervention but not meeting the major bleeding criteria.

## P2Y12 TREATMENT CHANGES

During the follow-up period, switch, or discontinuation in therapy, adherence, and persistence to the index P2Y12 APT were assessed.

A switch in therapy was defined as patients who discontinued use of a current P2Y12 DAPT for 30 days or more and replaced it with another antiplatelet drug.

Adherence is the degree to which a patient remains compliant to the prescribed medication and is operationally defined as the proportion of days covered (PDC) and calculated as the ratio of number of days P2Y12 APT was supplied and number of days in the follow-up period.^13^

Patients were considered to be adherent to DAPT if their PDCs were ≥80%.

## STATISTICAL ANALYSIS

Continuous variables were reported as mean±standard deviation, unless otherwise indicated.

We used the Analysis of Variance (ANOVA) test for intergroup comparison of various variables. A multivariate logistic regression model was used to identify the independent risk factors for CAD. The model included prespecified risk factors of old age, hypertension, diabetes mellitus, hyperlipidemia, current smoking, previous coronary artery disease, and a history of stroke.

A two-sided p value of <0.05 was considered statistically significant. Statistical analyses were performed using SPSS 22.0 (IBM, Armonk, NY, USA).

## RESULTS

### Baseline, laboratory, and angiographic characteristics

Among all patients, ticagrelor, prasugrel, and clopidogrel were prescribed to 29.5%, 4.3%, and 66.2% of patients, respectively. Table 1 shows baseline, laboratory, and angiographic characteristics. Clopidogrel-treated patients were older, female treated more with clopidogrel, ST elevation MI patients more frequent with less patients of higher NYHA functional class (NYHA ii-iv class), more patients deployed only single stent during percutaneous coronary intervention. Comorbidities were more frequent in ticagrelor and prasugrel group than clopidogrel group. In ticagrelor group more patients with diabetes 17(27.4%), CVA 3(4.8%), dyslipidaemia 15(24.2%), positive family history in 1(1.6%). Prasugrel group had more patients with hypertension (44.4%), smoking (66.7%), previous coronary artery disease.

**Table 1:**
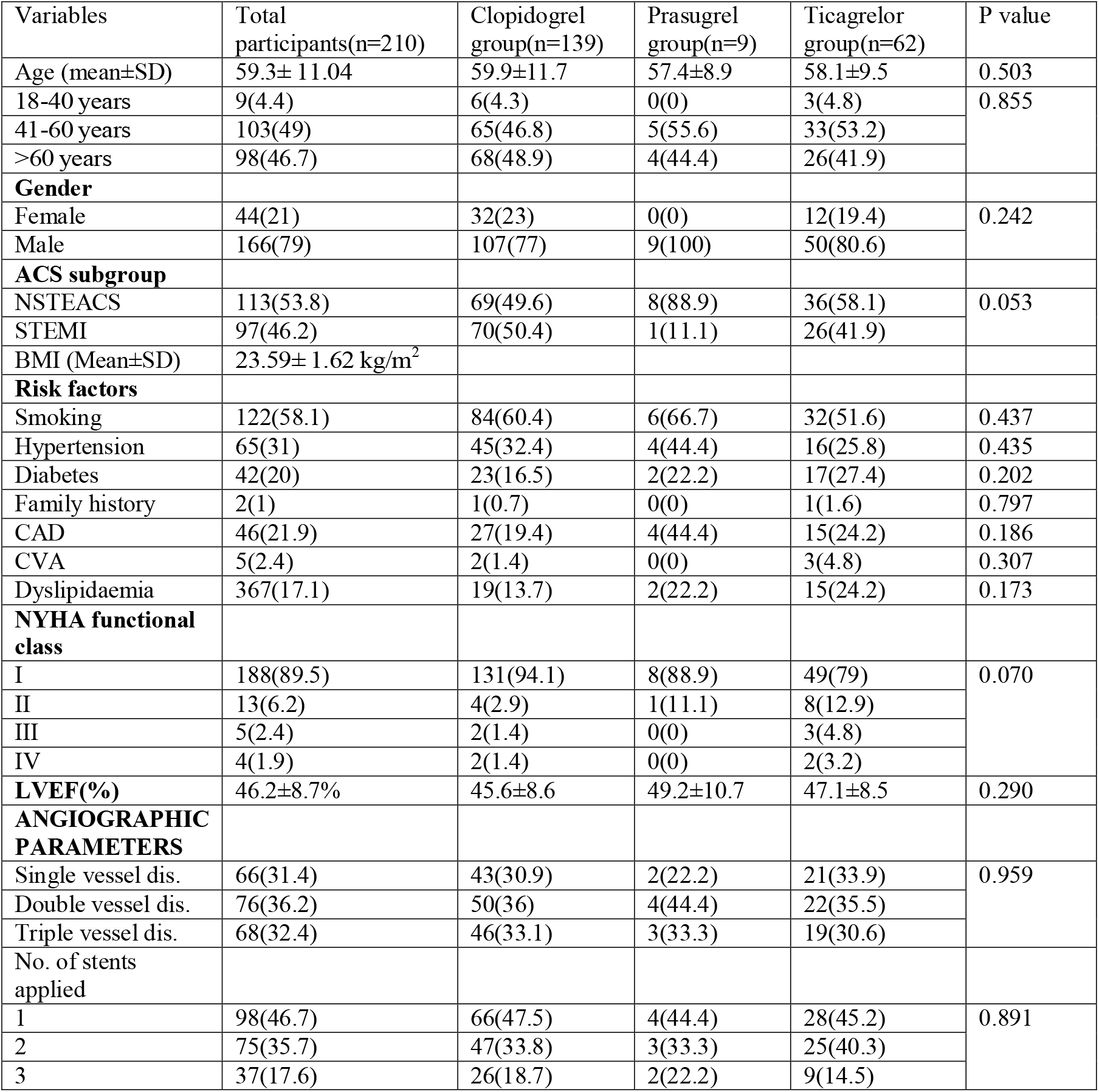
Sociodemographic, clinical and angiographic profile of study participants:

| Variables | Total participants(n=210) | Clopidogrel group(n=139) | Prasugrel group(n=9) | Ticagrelor group(n=62) | P value |
| --- | --- | --- | --- | --- | --- |
| Age (mean±SD) | 59.3± 11.04 | 59.9±11.7 | 57.4±8.9 | 58.1±9.5 | 0.503 |
| 18-40 years | 9(4.4) | 6(4.3) | 0(0) | 3(4.8) | 0.855 |
| 41-60 years | 103(49) | 65(46.8) | 5(55.6) | 33(53.2) |  |
| >60 years | 98(46.7) | 68(48.9) | 4(44.4) | 26(41.9) |  |
| **Gender** |  |  |  |  |  |
| Female | 44(21) | 32(23) | 0(0) | 12(19.4) | 0.242 |
| Male | 166(79) | 107(77) | 9(100) | 50(80.6) |  |
| **ACS subgroup** |  |  |  |  |  |
| NSTEACS | 113(53.8) | 69(49.6) | 8(88.9) | 36(58.1) | 0.053 |
| STEMI | 97(46.2) | 70(50.4) | 1(11.1) | 26(41.9) |  |
| BMI (Mean±SD) | 23.59± 1.62 kg/m<sup>2</sup> |  |  |  |  |
| **Risk factors** |  |  |  |  |  |
| Smoking | 122(58.1) | 84(60.4) | 6(66.7) | 32(51.6) | 0.437 |
| Hypertension | 65(31) | 45(32.4) | 4(44.4) | 16(25.8) | 0.435 |
| Diabetes | 42(20) | 23(16.5) | 2(22.2) | 17(27.4) | 0.202 |
| Family history | 2(1) | 1(0.7) | 0(0) | 1(1.6) | 0.797 |
| CAD | 46(21.9) | 27(19.4) | 4(44.4) | 15(24.2) | 0.186 |
| CVA | 5(2.4) | 2(1.4) | 0(0) | 3(4.8) | 0.307 |
| Dyslipidaemia | 367(17.1) | 19(13.7) | 2(22.2) | 15(24.2) | 0.173 |
| **NYHA functional class** |  |  |  |  |  |
| I | 188(89.5) | 131(94.1) | 8(88.9) | 49(79) | 0.070 |
| II | 13(6.2) | 4(2.9) | 1(11.1) | 8(12.9) |  |
| III | 5(2.4) | 2(1.4) | 0(0) | 3(4.8) |  |
| IV | 4(1.9) | 2(1.4) | 0(0) | 2(3.2) |  |
| **LVEF(%)** | 46.2±8.7% | 45.6±8.6 | 49.2±10.7 | 47.1±8.5 | 0.290 |
| **ANGIOGRAPHIC PARAMETERS** |  |  |  |  |  |
| Single vessel dis. | 66(31.4) | 43(30.9) | 2(22.2) | 21(33.9) | 0.959 |
| Double vessel dis. | 76(36.2) | 50(36) | 4(44.4) | 22(35.5) |  |
| Triple vessel dis. | 68(32.4) | 46(33.1) | 3(33.3) | 19(30.6) |  |
| No. of stents applied |  |  |  |  |  |
| 1 | 98(46.7) | 66(47.5) | 4(44.4) | 28(45.2) | 0.891 |
| 2 | 75(35.7) | 47(33.8) | 3(33.3) | 25(40.3) |  |
| 3 | 37(17.6) | 26(18.7) | 2(22.2) | 9(14.5) |  |

**Table 2:** Comparison of safety and efficacy and secondary objectives of study between participants received different P2Y12 inhibitors:

| Variables | Total participants(n=210) | Clopidogrel group(n=139) | Prasugrel group(n=9) | Ticagrelor group(n=62) | P value |
| --- | --- | --- | --- | --- | --- |
| All cause Death | 5(2.4) | 4(2.9) | 0(0) | 1(1.6) | 0.769 |
| ReMI | 4(1.9) | 2(1.4) | 0(0) | 2(3.2) | 0.633 |
| MACE | 9(4.2) | 6(4.3) | 0(0) | 3(4.8) | 0.529 |
| Bleeding | 6(2.9) | 3(2.2) | 1(11.1) | 2(3.2) | 0.289 |
| Switch over | 5(2.4) | 1(0.7) | 1(11.1) | 3(4.8) | **0.045** |
| Drug defaulter | 4(1.9) | 2(1.4) | 0(0) | 2(3.2) | 0.633 |

### Efficacy end points

After six month follow up, all-cause mortality was total 5 cases (2.4%) out of which 4 death (2.9%) occurred in clopidogrel group while 1 death (1.6%) occurred in ticagrelor group(p=0.76). no mortality was recorded in prasugrel group. Repeat myocardial infarction (Re MI) occurred in total 4 patients (1.9%) with 2 patients in clopidogrel and ticagrelor group each, and none from prasugrel group (p=0.633). No case of CVA reported during six month follow up.

### Safety end points

Bleeding occurred in total 6 patients (2.9%) out of which 3 patients (2.2%) in clopidogrel group, 1 patient (11.1%) in prasugrel group and 2 patients (3.2%) in ticagrelor group (p=0.29)). bleeding rates were higher in the ticagrelor or prasugrel groups than in the clopidogrel group, but no statistically significant differences existed in any bleeding between the ticagrelor, prasugrel and clopidogrel groups. The minor bleeding rates tended to increase only numerically in the ticagrelor or prasugrel groups compared with those in the clopidogrel group; however, there was no increase in the rates of major bleeding.

### Secondary objectives (drug defaulter and switch over rates)

During six months of follow up, total 5 patients (2.4%) switch over to another p2y12 inhibitor. 3 patients in ticagrelor group (4.8%) switch over to another p2y12 inhibitor, while one patient from each clopidogrel and prasugrel group switch over to another p2y12 group(p=0.045).

During this follow up period, a total of 4 patients (1.9%) were drug defaulter, out of which 2 were from clopidogrel group and two from ticagrelor group(p=0.622).

## DISCUSSION

This study was done in SMS hospital Jaipur to know about prescription rates and incidence of clinical outcomes, and to know drug defaulter and switch over rates in 210 consecutive ACS patients who underwent PCI after using ticagrelor, prasugrel, or clopidogrel. Prescription rates were ∼30.0% for ticagrelor, ∼4.0% for prasugrel, and 66.0% for clopidogrel. bleeding was observed in ∼3% of patients, but was observed more frequent in the ticagrelor or prasugrel groups; however, most of these bleeding events were minor. MACE occurred in ∼4% during a follow-up of 6 months.

Our hospital-based finding reflects the new P2Y12 inhibitors use in non-selected and real-world patients, rather than in a homogenous study population of randomized trials, and provides further information to guide the clinician in the choice of contemporary P2Y12 inhibitors. To achieve a better net clinical benefit in ACS patients following PCI, DAPT should be tailored to the individual patient’s ischemic and bleeding risks. Furthermore, ticagrelor or prasugrel have increased bleeding risks compared with clopidogrel. ^4-6,11,15^ So, this novel regimen of DAPT should be used in those with a high thrombotic or low bleeding risk.

Our findings indicate that ticagrelor or prasugrel based DAPT is not prescribed frequently to ACS patients undergoing PCI. Moreover, no statistically significant differences in all clinical outcomes between the use of ticagrelor or prasugrel over clopidogrel noted in our study. Although ticagrelor or prasugrel based DAPT is likely to improve clinical efficacies by decreasing the rate of cardiac death and preferred over clopidogrel based DAPT as per standard guidelines^1^, at a cost of higher bleeding tendency that consisted of mostly minor events. Findings of our study demonstrate that clopidogrel based DAPT can be preferred in our population safely over ticagrelor or prasugrel based DAPT with no significant increase in MACE or bleeding risk, which is more cost effective and already prescribed free of cost in institutional supply.

In our study, it was also observed that drug defaulter rates among various contemporary p2y12 groups was low and not significant during six months of follow up, but switch over to other p2y12 was significantly more in ticagrelor group than other groups. This signifies the effect of cost of drug and free supply in government hospitals effect adherence to a drug.

### Conclusions

As clopidogrel based DAPT was prescribed more in our centre because of free institutional supply and found no significant difference in clinical outcomes, although slight increase in bleeding risk in ticagrelor or prasugrel based DAPT, demonstrate clopidogrel based DAPT regime can be preferably prescribed safely in our population. Drug defaulter rates in first six month following PCI are low in all groups but more switch over from ticagrelor group to other groups significantly shows importance of low cost and free institutional supply in drug adherence.

### Limitations

First, this study was conducted in a single centre. Second, as sample size in our study was small, large sample size or a study involving regional or national registry sample population would be more informative statistically. Third, our study was prospective observational study, randomize control trial would be more informative. Fourth, follow up period in our study was six months, so information regarding long term clinical outcomes is not available.

## Data Availability

All data produced in the present study are available upon reasonable request to the authors

## Notes

### Competing Interest Statement

The authors have declared no competing interest.

### Funding Statement

This study did not receive any funding

### Author Declarations

Ethics committee of Sawai Man Singh Medical College And Attached Hospitals, Jaipur gave ethical approval for this work

